# Preferences for Tongue Swab versus Sputum Collection for Tuberculosis Testing: A Multi-Country Survey

**DOI:** 10.1101/2025.07.04.25330895

**Authors:** Kingsley Manoj Kumar, April Borkman, Ashley Kim, Rebecca Crowder, Bukola Ajide, Karla Alí-Francia, Masuzyo Chirwa, Louis Kamulegeya, Hien Le, Vu Ngoc Trung, Rouxjeane Venter, John Bimba, Devasahayam J. Christopher, Victoria Dalay, Nguyen Van Hung, Monde Muyoyeta, Lydia Nakiyingi, Nguyen Van Nhung, Grant Theron, Charles Yu, Carlos Zamudio-Fuertes, Julian Atim, Andrew D. Kerkhoff, Maria del Mar Castro Noriega, Payam Nahid, Claudia M. Denkinger, Adithya Cattamanchi, Susan E. Dorman, Nora West

**Author notes:** Co-first authors (Kingsley Manoj Kumar, April Borkman). Co-senior authors (Susan E. Dorman, Nora West).

## Abstract

**Background:** Sputum collection for tuberculosis (TB) diagnosis poses challenges for children, people living with HIV, and those who struggle with sputum production.

Tongue swab-based molecular testing offers a promising non-invasive alternative, but person-centered research on acceptability is limited.

**Methods:** We conducted a pragmatic survey across eight countries (Vietnam, Philippines, South Africa, Nigeria, Zambia, India, Uganda, Peru) among people with presumptive TB attending primary care facilities. Participants provided both tongue swab and sputum samples, then completed a 5-10 minute survey about their collection preferences.

**Results:** From October 2023 to July 2024, 1,297 participants were enrolled (median age 43 years, 45% female, 13% HIV-positive). Overall, 61% (95% CI: 58-64%) preferred tongue swab collection compared to 22% (95% CI: 20-25%) who preferred sputum collection and 17% (95% CI: 15-19%) with no preference. Preference for tongue swab was consistent across demographic and clinical subgroups, with country-level variation ranging from 47% in South Africa to 74% in Zambia and Nigeria.

**Conclusion:** Strong preference for tongue swab over sputum collection among individuals with presumptive TB supports this diagnostic innovation’s potential to overcome barriers to timely TB testing, particularly for populations struggling with sputum production.

---

Sputum collection is the specimen collection method for tuberculosis (TB) diagnosis. However, this approach poses challenges, particularly among children, people living with HIV (PLHIV), those who are severely ill, and others who experience difficulty producing sputum samples (Harries & Kumar, 2018). Tongue swab-based molecular testing is a promising, non-invasive method for TB detection that may provide advantages in speed, ease of use, and access compared to sputum-based molecular testing (Church et al., 2024).

Person-centered research on tongue swab-based TB testing is scarce (Church et al., 2024). This gap in understanding the perspectives of people affected by TB represents a critical limitation, as the successful implementation of any diagnostic innovation depends on acceptance by those impacted. Therefore, to inform future scale-up of tongue swab-based molecular testing, we conducted a pragmatic survey in eight countries among people with presumptive TB attending primary care facilities to ask about their experiences with and preferences for providing tongue swab compared to sputum samples.

Participants were enrolled from three parent studies evaluating novel tongue swab-based molecular diagnostics for TB: Rapid Research in Diagnostic Development for TB Network (R2D2 TB Network), Feasibility of Novel Diagnostics for TB in Endemic Countries (FEND-TB), and Assessing Diagnostics at Point-of Care for Tuberculosis (ADAPT).

All three studies enrolled people ≥12 years of age with presumptive TB based on having ≥2 weeks of new or worsening cough or a TB risk factor plus an abnormal WHO-endorsed TB screening test (R2D2 TB Network and ADAPT only) (Gupta-Wright et al., 2024). For this analysis, we included participants who provided sputum and tongue swab samples and completed the survey. The survey was administered by trained research staff and lasted approximately 5-10 minutes. Study procedures were approved by the institutional review boards (IRBs) of the University of California, San Francisco, the Heidelberg University Hospital, Rutgers University, the Foundation for Innovative New Diagnostics (Advarra), and by local IRBs at each enrollment site.

From October 2023 to July 2024, 1,297 participants were enrolled across eight countries: Vietnam, the Philippines, South Africa, Nigeria, Zambia, India, Uganda, and Peru. Median age was 43 years (IQR, 30–57 years), with the youngest participants in Nigeria (median, 34 years) and the oldest in Vietnam (median, 56 years). Overall, 45% of participants were female (range: 31% in Uganda to 60% in Peru), 13% were living with HIV (range: 0% in the Philippines to 51% in Uganda), 17% reported a history of diabetes mellitus (range: 3% in Uganda to 42% in Nigeria), 23% reported current smoking (range: 5% in Nigeria to 45% in South Africa), 20% reported prior TB treatment (range: 11% in Nigeria to 39% in the Philippines) and 95% were enrolled based on having cough for ≥2 weeks.

Among all participants, 61% (95% CI: 58-64%) preferred tongue swab collection, compared to 22% (95% CI: 20–25%) who preferred sputum collection and 17% (95% CI: 15-19%) who had no preference. Tongue swab collection was preferred by 65% of females (95% CI: 61-69%) and 58% of males (95% CI: 54-61%). Preference for tongue swab collection was highest among participants aged 18–29 (71%, 95% CI: 66-77%) and lower among those aged 45-59 years (56%, 95% CI: 50-61%). Across countries, preference for tongue swab collection was highest in Zambia (74%, 95% CI: 65-81%) and Nigeria (74%, 95% CI: 66–80%), and lowest in South Africa (47%, 95% CI: 41-54%) (Figure 1).

**Figure 1.**
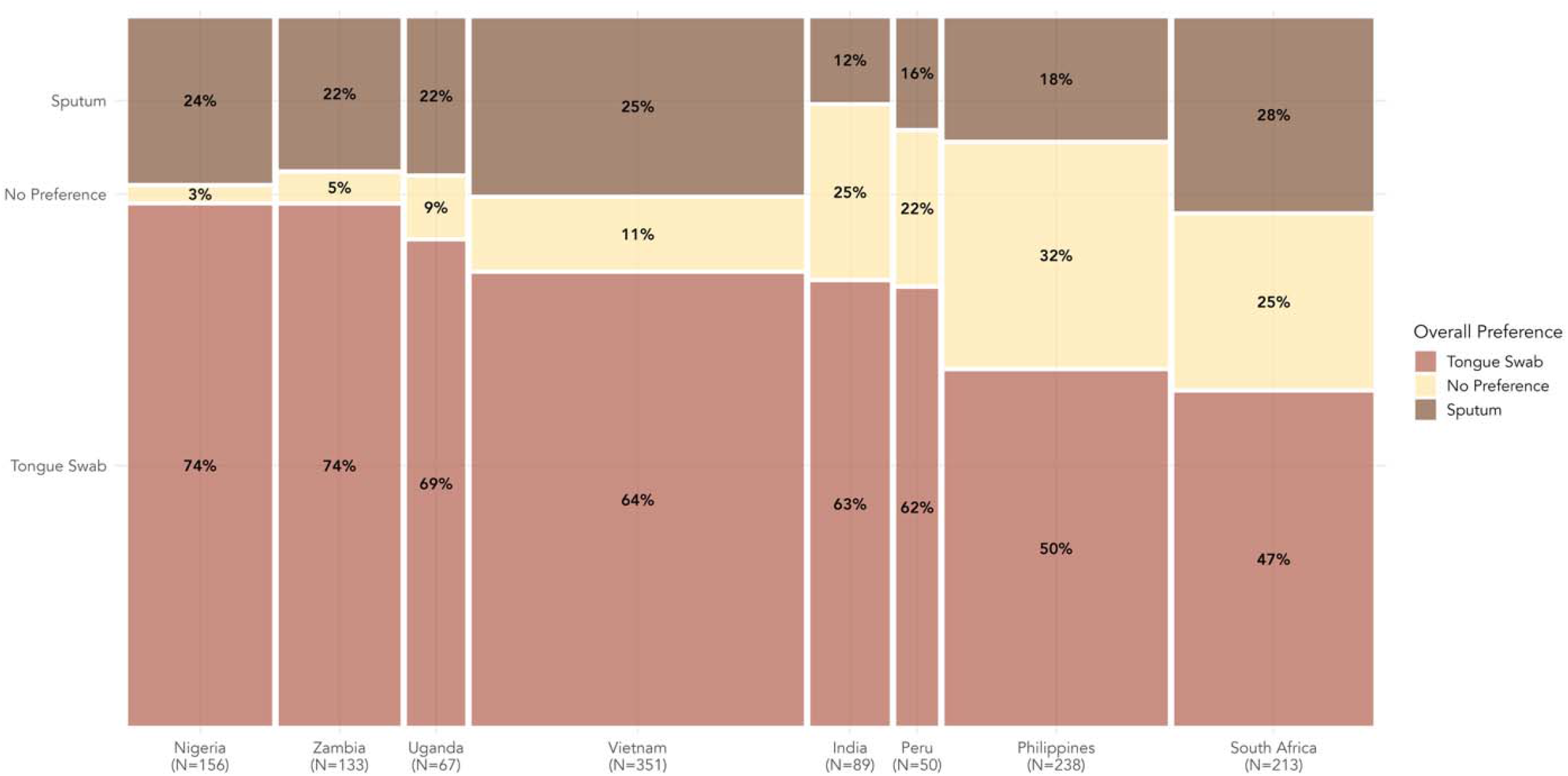
Mosaic plot with preference for tongue swab and sputum sample by country and sample size. The width of each bar represents the relative number of participants per country, and the height of each segment represents the proportion within each country indicating a given preference (tongue swab, sputum, or no preference).

Clinical factors were not associated with a substantial difference in preference for tongue swab vs. sputum collection. Participants living with vs. without HIV both preferred tongue swab collection (62% vs. 61%, p=0.18). Participants who provided expectorated and induced sputum samples both preferred tongue swab collection (61% and 62%, respectively), although there was a higher preference for sputum collection among those who could expectorate sputum (24% vs. 13%, p=0.001).

Our findings suggest a strong preference for tongue swab over sputum collection among individuals with presumptive TB, with over 60% of participants preferring tongue swab collection. This was consistent across demographic and clinical subgroups, suggesting broad acceptability of this diagnostic modality.

Recent research has shown promising diagnostic performance of tongue-swab-based molecular tests. Steadman et al. found that tongue swab-based molecular testing could achieve sensitivity of 93% and specificity of 99% compared to a microbiological reference standard when using optimized processing protocols paired with qPCR (Steadman et al., 2024). Similar results have now been demonstrated with novel semi-automated platforms designed for swab-based molecular testing (Steadman et al., 2025). Our findings complement studies on diagnostic accuracy by offering a person-focused perspective on acceptability. While we found an overall preference for tongue swab collection among people with presumptive TB, country-level variations in preference (47% in South Africa to 74% in Zambia and Nigeria) warrant further investigation, and indicate the need to potentially provide individuals with options for sample collection. These differences may reflect varying cultural contexts, prior exposure to different diagnostic methods, how tongue swab collection was implemented across sites, or a lack of a strong preference between the two sample types, as seen in some countries (up to 32% in the Philippines) and prior TB test preference studies (del Mar Castro et al., 2024; Shah et al., 2024). Future research should explore these contextual factors to optimize implementation strategies.

Our study had several strengths, including enrollment of people with presumptive TB from eight high TB burden countries, having participants’ preferences informed by first experiencing both sample collection types, and having the survey conducted by a different person than the one who collected the samples, minimizing interviewer and social desirability bias. However, our research has some limitations. First, participants provided tongue swabs before sputum samples, which may have influenced their reported preferences. Second, our study population was limited to people with TB symptoms and/or risk factors seeking care at health facilities. Despite these limitations, our findings provide compelling evidence that tongue swab collection is preferred over traditional sputum collection across diverse populations and settings.

In summary, when combined with the emerging data on its diagnostic accuracy, our findings further support tongue swab-based molecular testing as a promising diagnostic innovation that could help overcome barriers to timely TB testing, particularly for populations that struggle with sputum production. This diagnostic innovation aligns with global priorities to end the TB epidemic by improving testing accessibility through expanded testing methods. Although further work is needed to assess the implementation of tongue swab testing and its yield in diverse real-world settings, our findings provide strong support for its role in the evaluation of presumptive TB patients.

## Data Availability

All data produced in the present work are contained in the manuscript.

## Disclosure

The author reports no conflicts of interest in this communication.

## Funding

ADAPT is funded by the Supporting, Mobilizing, and Accelerating Research for Tuberculosis Elimination (SMART4TB) Consortium, made possible by the generous support of the American people through the United States Agency for International Development (USAID) and implemented under cooperative agreement number 7200AA20CA00005. The FEND-TB Consortium is funded by the National Institutes of Health (NIH) (funding reference number U01AI152084). The R2D2 Network is funded by the NIH (funding reference number U01AI152087). The communication is the authors’ sole responsibility and does not necessarily reflect the views of NIH, USAID, the US Government, or consortium collaborators or members.

## Notes

### Competing Interest Statement

The authors have declared no competing interest.

### Author Declarations

Ethics committees/IRBs of the University of California, San Francisco, the Heidelberg University Hospital, Rutgers University, the Foundation for Innovative New Diagnostics (Advarra), and by local IRBs at each enrollment site gave ethical approval for this work.

